# The influence of acute dietary nitrate supplementation on skeletal muscle fatigue and recovery in older women

**DOI:** 10.1101/2023.02.15.23285957

**Authors:** William S. Zoughaib, Richard L. Hoffman, Brandon A. Yates, Ranjani N. Moorthi, Kenneth Lim, Andrew R. Coggan

**Author notes:** **Corresponding Author:** Andrew R. Coggan, PhD, Department of Kinesiology, Indiana University Purdue University Indianapolis, IF 101C, 250 University Boulevard, Indianapolis, IN 46112.

## Abstract

Older individuals fatigue more rapidly during, and recover more slowly from, dynamic exercise. Women are particularly vulnerable to these deleterious effects of aging, which increases their risk of falling. We have shown that dietary nitrate (NO_3_^-^), a source of nitric oxide (NO) via the NO_3_^-^ → nitrite (NO_2_^-^) → NO pathway, enhances muscle speed and power in older individuals in the non-fatigued state; however, it is unclear if it reduces fatigability and/or improves recoverability in this population. Using a double-blind, placebo-controlled, crossover design, we studied 18 older (age 70 ± 4 y) women who were administered an acute dose of beetroot juice (BRJ) containing either 15.6±3.6 or <0.05 mmol of NO_3_^-^. Blood samples were drawn throughout each ∼3 h visit for plasma NO_3_^-^ and NO_2_^-^ analysis. Peak torque was measured during, and periodically for 10 min after, 50 maximal knee extensions performed at 3.14 rad/s on an isokinetic dynamometer. Ingestion of NO_3_^-^-containing BRJ increased plasma NO_3_^-^ and NO_2_^-^ concentrations by 21±8 and 4±4 fold, respectively. However, there were no differences in muscle fatigue or recovery. Dietary NO_3_^-^ increases plasma NO_3_^-^ and NO_2_^-^ concentrations but does not reduce fatigability during or enhance recoverability after high intensity exercise in older women.

## INTRODUCTION

As humans age, there are detrimental changes in physical function, which are due in large part to changes in the properties of skeletal muscle. These include decreases in the maximal force, speed, and power of muscle contraction [Larsson et al. 2019], as well as reductions in mitochondrial respiratory capacity [Ferri et al. 2020] and in blood flow during exercise [Ridout et al. 2010; Casey et al. 2015]. The latter lead to a greater disturbance of cellular energetics during activity [Coggan et al. 1993; Lewsey et al. 2020], which along with the overall slowing of contractile properties result in increased fatigability during dynamic exercise [Christie et al. 2012], especially at higher movement velocities [Dalton et al. 2010; Callahan and Kent-Braun 2011]. The rate of recovery from fatiguing exercise, i.e., recoverability, is also diminished in older individuals [Schwendner et al. 1997; Yoon et al. 2012]. Since most activities of daily living are intermittent, not continuous, in nature, the latter may be of particular practical and clinical relevance. Women are especially susceptible to the deleterious effects of aging on physical function [Murtaugh and Hubert 2004; Freedman et al. 2016], with increased fatigability and reduced recoverability of muscle predisposing them to falls [Schwendner et al. 1997] and an increased risk hospitalization and even institutionalization.

Dietary nitrate (NO_3_^-^) supplementation may provide a viable means of offsetting some of these detrimental age-related changes in skeletal muscle function. Upon ingestion and concentration/secretion by the salivary glands, NO_3_^-^ can be reduced by oral bacteria to form nitrite (NO_2_^-^), which once absorbed can in turn be reduced by, e.g., deoxyhemoglobin to generate nitric oxide (NO) [Lundberg and Weitzberg 2009]. Given the importance of NO in modulating muscle function, including contractility, mitochondrial respiration, blood flow, etc. [Stamler and Meissner 2001], numerous studies in recent years have therefore assessed the effects of dietary NO_3_^-^ (usually in the form of beetroot juice (BRJ)) on exercise capacity. As reviewed elsewhere, such studies have demonstrated that acute or short-term dietary NO_3_^-^ supplementation can improve the inherent contractile properties of muscle (Coggan et al. 2021; Esen et al. 2022] as well as enhance performance during high intensity dynamic exercise [Silva et al. 2022], at least/especially during open-ended tests performed by non-athletes. Increasing NO bioavailability via NO_3_^-^ intake could therefore potentially ameliorate at least some of the negative effects of aging described above. Indeed, we recently found that acute ingestion of BRJ containing 13.4 mmol of NO_3_^-^ significantly increased maximal knee extensor speed and power in twelve 71 y old men and women [Coggan et al. 2020a]. However, there were no changes in fatigability during an “all-out”, 50 contraction (∼1 min) fatigue test conducted at 3.14 rad/s on an isokinetic dynamometer. The latter finding could be due to the numerous factors likely contributing to muscle fatigue under such conditions (e.g., changes in high energy phosphate and/or H^+^ concentrations, altered Ca^2+^ handling kinetics, impaired excitation-contraction coupling, etc. [Kent-Braun et al. 2012]), only some of which might be influenced by increased NO bioavailability. However, they could also be due to the relatively small sample size, or to the fact that we studied older men as well as women when we have found that women appear to be more likely to benefit from NO_3_^-^ supplementation [Coggan et al. 2018b].

Although dietary NO_3_^-^ may not (or may) enhance muscle fatigue resistance in older persons, it might still improve the rate of recovery from fatiguing activities. As a potent vasodilator, NO could increase muscle blood flow post-exercise, when intramuscular pressure is no longer elevated by contractions. This might speed the “washout” of fatigue-related metabolites such as H^+^ from muscle [Kemp et al. 1997]. Furthermore, by delivering more O_2_, higher post-exercise blood flow could increase the rate of mitochondrial ATP synthesis, thus resulting in more rapid restoration of high energy phosphate levels [Layec et al. 2013] and hence muscle function [Jansson et al. 1990]. In fact, two studies of healthy young men and women found that under hypoxic conditions dietary NO_3_^-^ accelerated phosphocreatine (PCr) recovery kinetics following high intensity knee extensor exercise [Vanhatalo et al. 2011, 2014], indicative of increased O_2_ delivery via enhanced blood flow and/or improved mitochondrial coupling. However, NO_3_^-^ingestion had no effect on PCr recovery in healthy older men and women studied under normoxic conditions [Kelly et al. 2013]. Thus, whether dietary NO_3_^-^ supplementation might augment recovery of muscle function following fatiguing exercise in older persons is unclear.

The purpose of the present study was to test the hypothesis that acute dietary NO_3_^-^ supplementation would reduce fatigability and/or improve recoverability of muscle specifically in older women. If so, this could have broad implications for this population of individuals, who are particularly vulnerable to the deleterious effects of reductions in muscle function with aging.

## MATERIALS AND METHODS

### Participants

A total of 18 community-dwelling women with a mean age, height, weight, and body mass index of 70±4 y, 1.65±0.06 m, 67.1±9.5 kg, and 24.6±3.2 kg/m^2^, respectively, completed this registered clinical trial (NCT03595774). These women were studied after screening a total of 143 potential participants, of whom n=98 were deemed ineligible based on a phone screen and review of their medical records, n=19 declined to participate or were lost to follow-up, and n=8 failed to meet inclusion criteria based on an initial visit to the University Hospital Clinical Research Center (CRC). During this visit, potential participants underwent a health history, physical examination, resting ECG, and phlebotomy (CBC, CMP, fasting insulin, and lipids), and also practiced the isokinetic dynamometry testing protocol (see below).

Individuals were excluded if they were not between the ages of 65 and 79 y, were unable to provide informed consent, were currently smokers, pregnant, or lactating, were taking proton pump inhibitors, antacids, or xanthine oxidoreductase inhibitors, or had a history of major metabolic (thyroid disorders, type I or II diabetes), cardiovascular (e.g., moderate or severe valvular disease, myocardial/pericardial disease, stage II or greater hypertension, heart failure, myocardial infarction/ischemia), renal (eGFR <60 mL/min/1.73 m^2^, or 61-90 ml/min/1.73 m^2^ and albumin:creatine ratio > 30), neuromuscular (e.g., cervical spondylotic radiculomyelopathy, lumbar spondylosis, amyotrophic lateral sclerosis, Guillain-Barre syndrome, acquired demyelinating polyneuropathies), or liver (e.g., SGOT/SGPT >2x normal) diseases, were anemic (hematocrit < 30%), or had any other contraindications to vigorous exercise. Other exclusion criteria included whether the participant was on hormone replacement therapy or used phosphodiesterase inhibitors, since these can respectively diminish [Obach et al. 2004] or potentiate [Webb et al. 1999] the effects of dietary NO_3_^-^. All other participants were included. The Human Subjects Office at Indiana University approved this study and written informed consent was obtained from each individual.

### Experimental Design and Protocol

Starting approximately 1 wk after completion of the initial screening visit, eligible participants were studied using a randomized, double-blind, placebo-controlled crossover design. Participants were asked to avoid foods high in NO_3_^-^ (e.g., spinach, arugula, beets) for 24 h and alcohol, caffeine, or food for 12 h before testing. Otherwise, no dietary restrictions were imposed. Upon arrival at the CRC, an intravenous catheter was placed and a baseline blood sample taken for future analysis of plasma NO_3_^-^ and NO_2_^-^ concentrations as described below. The participant then ingested 140 mL of a concentrated beetroot juice (BRJ) supplement (Beet it Sport, James White Drinks, Ipswich, UK) containing (based on direct measurement) either 15.6±3.6 or <0.05 mmol of NO_3_^-^. Extraction of NO_3_^-^ from BRJ to create the latter placebo was performed by the manufacturer using a highly selective anion exchange resin. After 1 and 2 h of quiet rest, additional blood samples were collected, after which muscle function testing was performed as described below. Ten minutes later, a final blood sample was obtained, after which participants were fed lunch then released. They then returned to the CRC after a 1 wk washout period and repeated the experiment, this time ingesting the opposite BRJ supplement than before. Participants therefore made a total of three study visits, i.e., initial screening, placebo BRJ, NO_3_^-^-containing BRJ, with the latter two in random order.

### Measurement of Muscle Function

Fatigability and recoverability of the knee extensor muscles were assessed using isokinetic dynamometry (Biodex System 4 Pro, Biodex Medical Systems, Shirley, NY). The femoral condyle of the participant’s dominant leg was aligned with the dynamometer’s axis of rotation while the lower leg, thigh, waist, and torso were securely restrained with straps to prevent any extraneous movement. The participant first performed three maximal knee extensions at each of five velocities (i.e., 0, 1.57, 3.14, 4.71, and 6.28 rad/s), with 120 s of rest between sets, to determine their peak torque-velocity relationship (data reported separately). After an additional 120 s of rest, they then performed 50 maximal knee extensions at a velocity of 3.14 rad/s. The velocity of knee flexion was set to 6.28 rad/s, allowing the participant to freely reposition their leg against minimal resistance between repetitions. Participants were instructed to go “all-out” during this fatigue test, i.e., to extend their leg as quickly and as forcefully as possible during each repetition and to not pace themselves in an attempt to maintain torque. Strong verbal encouragement was provided throughout each test. After completion of the fatigue test, participants performed sets of three additional maximal knee extensions at 3.14 rad/s after 30, 60, 150, 300, and 600 s (nominally) of recovery.

Data from the isokinetic dynamometer were processed by windowing, filtering, and smoothing as previously described [Coggan et al. 2015]. Fatigability was assessed based on the average torque, total work, work during the first and last 1/3^rd^ of the fatigue test, and the ratio of the latter as reported by the manufacturer’s software. The torque data were also exported and analyzed to determine the peak torque during each of the 50 contractions. Recoverability was assessed based on the degree of restoration of peak torque at the different time points following the fatigue test. Because it was not possible to measure peak torque at identical times in every participant, a monoexponential function was also fit to the peak torque-recovery duration data (including the final value from the fatigue test) for each individual to determine their time constant (τ) and half-life (=ln(2)/τ) of torque recovery:

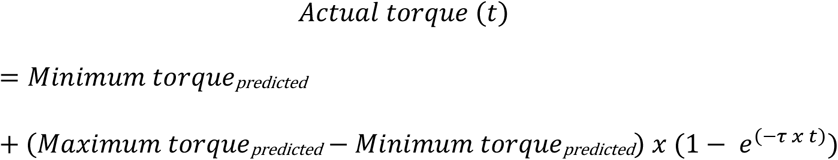

where t = the exact time after the 50 contraction fatigue test and torque is expressed as a percentage of the maximum torque observed at the start of the fatigue test (which almost always occurred during the 2^nd^ or 3^rd^ of the 50 knee extensions).

### Measurement of Plasma NO_3_^-^ and NO_2_^-^

A dedicated high-performance liquid chromatography system (ENO-30, Eicom USA, San Diego, CA) was used to measure plasma NO_3_^-^ and NO_2_^-^ concentrations as previously described in detail [Coggan et al. 2018a]. Briefly, 25 μL of thawed plasma was mixed 1:1 with methanol, centrifuged, and then a 10 μL aliquot of the protein-poor supernatant was injected into the HPLC. Plasma NO_3_^-^ and NO_2_^-^concentrations were determined from calibration curves generated using NIST-traceable standard solutions.

### Statistical Analysis

Data were analyzed using GraphPad Prism version 9.3.1 (GraphPad Software, La Jolla, CA). The normality of data distribution was tested using the D’Agostino-Pearson omnibus test. Plasma NO_3_^-^ and NO_2_^-^ and peak torque data were compared between trials using two-way (treatment x time or treatment x contraction number) repeated measures ANOVA. Post-hoc testing was performed using the Holm-Šidák multiple comparison procedure. Fatigue test summary statistics (e.g., average torque) and monoexponential recovery curve fit parameters were compared between trials using paired t-tests. Multiplicity-corrected P values less than 0.05 were considered significant. Deidentified data are available from the authors upon request.

## RESULTS

### Plasma NO_3_^-^ and NO_2_^-^ levels

There were no significant changes in plasma NO_3_^-^ or NO_2_^-^ levels following ingestion of the NO_3_^-^-depleted BRJ placebo (Table 1). In contrast, plasma NO_3_^-^ and NO_2_^-^ concentrations were significantly elevated 1 and 2 h post-ingestion of NO_3_^-^-containing BRJ, as well as 10 min after completion of the exercise testing (approximately 2.5 h post-ingestion).

**Table 1.** Effects of NO_3_^-^ supplementation on plasma NO_3_^-^ and NO_2_^-^ concentrations.

|  |  | Time of measurement |  |  |  |
| --- | --- | --- | --- | --- | --- |
|  |  | Pre | 1 h post | 2 h post | 10 min post |
|  |  | <u>ingestion</u> | <u>ingestion</u> | <u>ingestion</u> | <u>exercise</u> |
| Plasma [NO<sub>3</sub><sup>-</sup>]<br>(μmol/L) | Placebo | 47.7 | 47.7 | 51.9 | 51.1 |
|  |  | ±30.3 | ±23.5 | ±25.9 | ±24.4 |
|  | Nitrate | 42.6 | 622.9 | 751.3 | 735.2 |
|  |  | ±17.7 | ±227.6 | ±156.9 | ±158.2 |
| P value |  | 0.312 | 1.20 x 10<sup>-7</sup> | 3.35 x 10<sup>-10</sup> | 5.60 x 10<sup>-10</sup> |
| Plasma [NO<sub>2</sub><sup>-</sup>]<br>(μmol/L) | Placebo | 0.343 | 0.363 | 0.387 | 0.411 |
|  |  | ±0.230 | ±0.286 | ±0.358 | ±0.330 |
|  | Nitrate | 0.376 | 0.786 | 0.968 | 1.209 |
|  |  | ±0.286 | ±0.783 | ±0.803 | ±0.883 |
| P value |  | 0.404 | 2.28 x 10<sup>-2</sup> | 4.50 x 10<sup>-3</sup> | 2.45 x 10<sup>-3</sup> |

Values are mean±S.D. for n=18.

### Fatigability

There were no significant differences between placebo and NO_3_^-^ trials in the time required to complete the 50 contraction fatigue test (i.e., 62±6 vs. 64±9 s; P=0.153) or in the total time spent extending (33±2 vs. 34±2 s; P=0.078) or flexing (29±4 vs. 30±7 s; P=0.384) the leg. There were also no significant differences between trials in the average torque, total work, work during the first 1/3^rd^ or last 1/3^rd^ of the test, or their ratio (Table 2). Finally, analysis of the raw data to determine the peak torque during each of the 50 contractions also revealed no significant treatment (P=0.851) or interaction (P=0.963) effects (Fig. 1).

**Table 2.** Fatigue test summary results.

| <u>Trial</u> | Average<br>Torque<br>(Nm/kg) | Total<br>work<br>(J/kg) | Work<br>first 1/3<sup>rd</sup><br>(J/kg) | Work<br>last 1/3<sup>rd</sup><br>(J/kg) | Fatigue<br>index<br>(%) |
| --- | --- | --- | --- | --- | --- |
| Placebo | 0.57<br>±0.10 | 34.6<br>±8.0 | 15.7<br>±8.0 | 8.0<br>±2.1 | 50.8<br>±4.8 |
| Nitrate | 0.57<br>±0.10 | 35.3<br>±8.2 | 16.0<br>±3.7 | 8.2<br>±2.1 | 51.4<br>±5.8 |
| P value | 0.969 | 0.260 | 0.204 | 0.229 | 0.593 |

Values are mean±S.D. for n=18.

**Figure 1.**
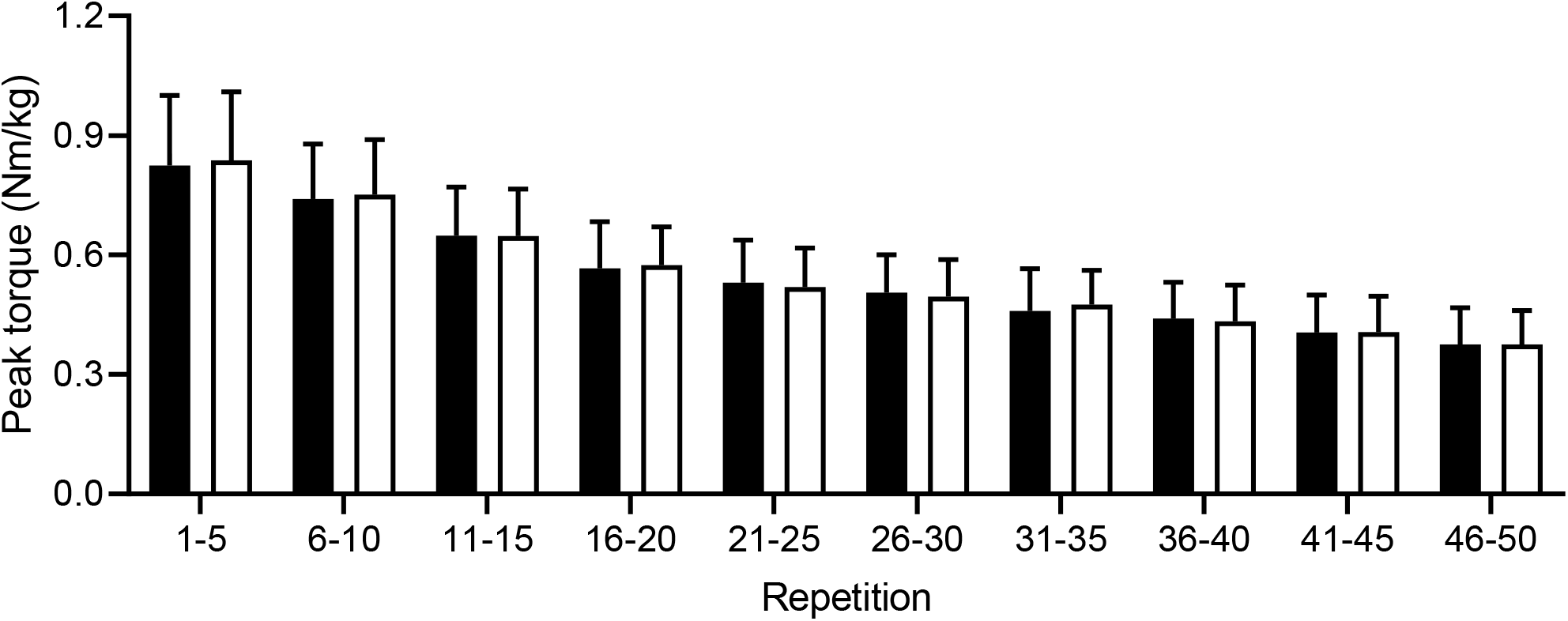
Knee extensor peak torque during the fatigue test. Note that although 5 repetition averages are presented for clarity, statistical analysis was performed using the peak torque from each of the 50 repetitions. Values are mean±S.D. for n=18. No significant differences were observed.

### Recoverability

Peak torque was measured after 35±3, 64±5, 159±19, 309±20, and 603±4 s of recovery in the placebo trial and after 34±3, 65±2, 154±3, 302±4, and 603±7 s of recovery in the NO_3_^-^ trial (treatment effect, P=0.161; interaction effect, P=0.239). No significant differences due to NO_3_^-^ ingestion were observed, regardless of whether torque was expressed relative to body mass (treatment effect, P=0.326; interaction effect, P=0.281) or as a percentage of the non-fatigued baseline (treatment effect, P=0.275; interaction effect, P=0.500) (Fig. 2). There were also no differences in the predicted minimum or maximum torque or time course of recovery of torque determined via exponential curve fitting (Table 3).

**Table 3.** Curve fit parameters for recovery of isokinetic torque at 3.14 rad/s.

| <u>Trial</u> | Minimum torque<br><u>(% of non-fatigued)</u> | Maximum torque<br><u>(% of non-fatigued)</u> | Time constant<br><u>(s<sup>-1</sup>)</u> | Half-life<br><u>(s)</u> | S.E.E.<br><u>(%)</u> |
| --- | --- | --- | --- | --- | --- |
| Placebo | 45.6 | 97.8 | 0.01515 | 72.4 | 4.3 |
|  | ±7.9 | ±8.0 | ±0.00898 | ±82.0 | ±1.2 |
| Nitrate | 45.8 | 97.4 | 0.01457 | 58.1 | 4.7 |
|  | ±6.2 | ±5.5 | ±0.00671 | ±26.7 | ±1.6 |
| P value | 0.882 | 0.856 | 0.785 | 0.506 | 0.336 |

Values are mean±S.D. for n=18.

**Figure 2.**
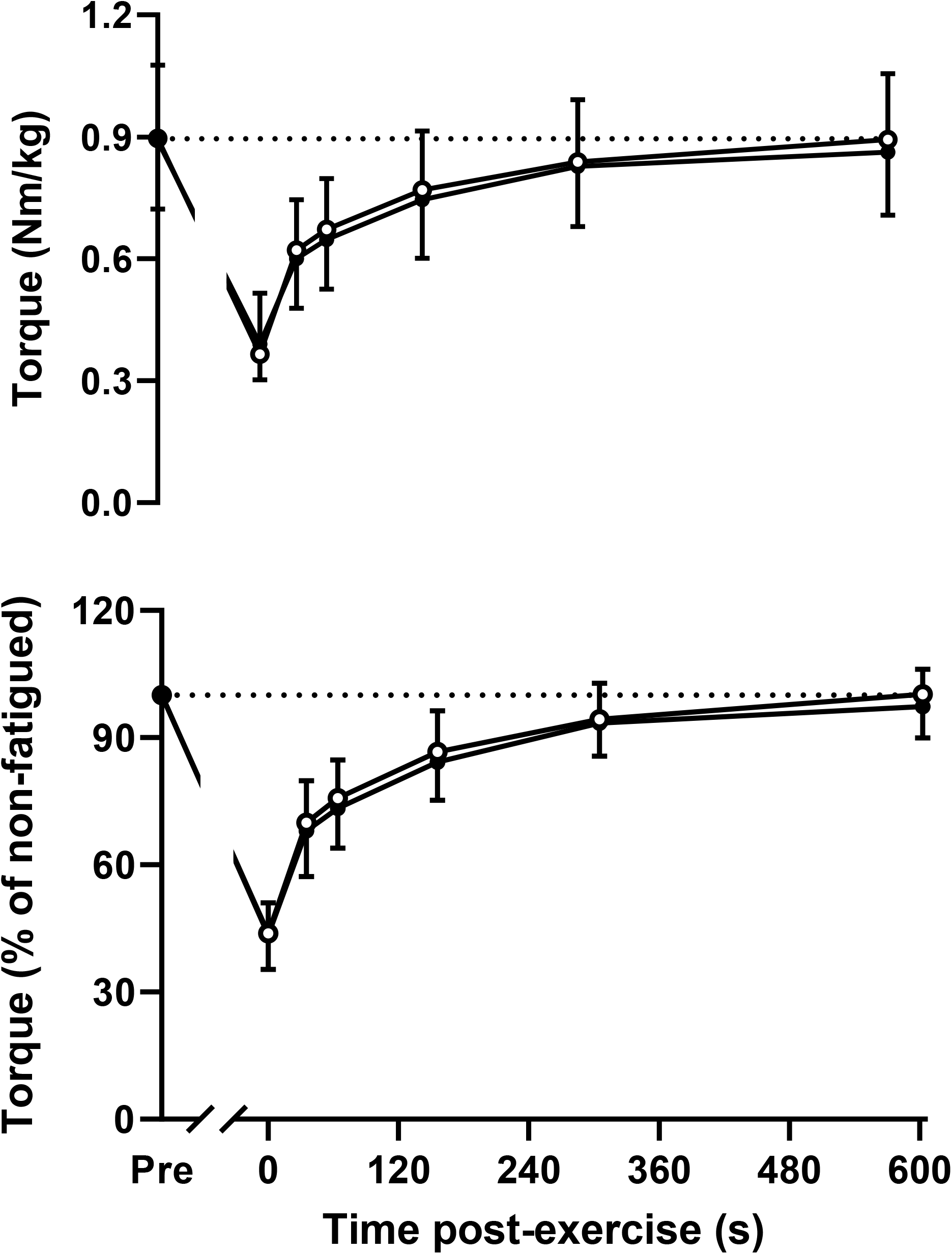
Recovery of knee extensor peak torque following the fatigue test. Data are expressed either relative to body mass (*top panel*) or as a percentage of the maximum non-fatigued value (*bottom panel*). Values are mean±S.D. for n=18. No significant differences were observed.

## DISCUSSION

Aging results in more rapid fatigue during, and slower recovery from, dynamic exercise. Older women in particular are prone to these negative effects of aging, which increases their risk of falls. The purpose of the present study was to determine whether acute dietary NO_3_^-^ intake might ameliorate these age-related changes in muscle function. However, despite producing large changes in plasma NO_3_^-^ and NO_2_^-^ levels, and hence presumably in NO bioavailability, we found no effect of such supplementation on either the fatigability or recoverability of muscle in this population.

In the present study, the ingestion of BRJ containing 15.6±3.6 mmol of NO_3_^-^ increased the plasma concentration 21±8-fold, or to an individual peak of 783±139 μmol/L above baseline. Assuming essentially complete absorption, this corresponds to a volume of distribution of 20.1±3.3 L, which agrees exactly with our recent novel pharmacokinetic modeling study of the combined NO_3_^-^/NO_2_^-^ system [Coggan et al. 2020b]. Plasma NO_2_^-^ concentrations also rose, albeit to a lesser extent, i.e., 4±4-fold, due to the larger volume of distribution and almost 70-fold more rapid clearance of the latter and the fact that only 3% of ingested NO_3_^-^ is converted to NO_2_^-^ [Coggan et al. 2020b]. These changes in plasma NO_3_^-^ and NO_2_^-^ are consistent with numerous previous studies of dietary NO_3_^-^ supplementation that have employed a similar dose [e.g., Jonvik et al. 2018]. We did not measure breath NO levels as we have in our previous studies of NO_3_^-^ supplementation in older individuals [Coggan et al. 2020a; Gallardo et al., 2021], but based on such data the latter likely would have increased by at least 100%. It therefore seems reasonable to conclude that the ingestion of NO_3_^-^-containing BRJ was effective in increasing whole-body NO bioavailability.

Despite the above, acute NO_3_^-^ supplementation has no impact on muscle fatigability during a 50 contraction (∼60 s) isokinetic knee extensor exercise test. The present results corroborate and extend our prior study of a smaller group of older women and men combined [Coggan et al. 2020a]. Although direct comparisons are challenging due to potential differences between small muscle mass and whole-body exercise, they are also consistent with those of prior studies examining the effects of NO_3_^-^ ingestion on, e.g., 30 s Wingate test performance in younger, healthier individuals [Rimer et al. 2016; Jonvik et al. 2018]. Indeed, a recent meta-analysis by Silva et al. [2022] concluded that dietary NO_3_^-^ is effective at improving performance during dynamic exercise lasting 2-10 min, but not during shorter or longer efforts. In contrast, Kadach et al. [2023] recently found that NO_3_^-^ supplementation increased muscle torque throughout the first 90 s of a 300 s bout of repeated maximal isometric knee extensions (3 s contraction, 2 s rest). Notably, however, numerous other investigations have failed to demonstrate any effect of dietary NO_3_^-^ on maximal isometric torque [Esen et al. 2022], making the study by Kadach et al. [2023] an exception. Regardless, based on the present as well as our previous results, it seems clear that although acute ingestion of NO_3_^-^ enhances maximal muscle speed and hence power in older individuals [Coggan et al. 2020a; Gallardo et al. 2021], it does not alter fatigability during intense contractile activity in older persons.

A novel aspect of the present investigation was determination of the effects of dietary NO_3_^-^ on recovery of muscle contractile function following fatiguing exercise. Specifically, we hypothesized that NO_3_^-^ supplementation might enhance recovery of muscle torque by magnifying post-exercise hyperemia. Reductions in intramuscular pO_2_ and/or pH during exercise should accelerate conversion of NO_3_^-^/NO_2_^-^ to NO [Lundberg and Weitzberg 2009], which in turn could lead to greater vasodilation and higher blood flow following exercise, once intramuscular pressure decreases. If so, this could result in more rapid restoration of muscle pH and/or high energy phosphate levels, and hence muscle function. Indeed, NO_3_^-^ intake has been previously reported to increase PCr recovery kinetics in healthy young men and women studied in hypoxia [Vanhatalo et al. 2011, 2014], but not in younger [Vanhatalo et al. 2014] or older [Kelly et al. 2013] individuals studied in normoxia. Although we did not measure changes in muscle pH and/or high energy phosphate levels in the older women in the present study, acute NO_3_^-^ ingestion had no effect on the recovery of muscle torque following fatiguing exercise. Thus, either any increase in post-exercise blood flow was insufficient to result in more rapid restoration of function, or increased flow simply did not improve recovery, reflecting the likely multifactorial nature of fatigue under the present conditions [Kent-Braun et al., 2012].

There are a number of strengths to the present study, including a relatively large sample size, careful screening of participants to avoid confounding the effects of aging with those due to chronic disease, and the rigorous evaluation of muscle fatigability/recoverability during/after high intensity dynamic exercise. There are, however, a number of limitations as well. Specifically, as indicated above we did not collect any mechanistic measurements (e.g., changes in blood flow, PCr kinetics, etc.) that could have provided further insight into the effects of acute NO_3_^-^ intake on skeletal muscle physiology in this population, even if function was unchanged. It is also possible that we might have obtained different results had we utilized a different dose of NO_3_^-^, had studied the participants after multiple days of supplementation, and/or had studied men instead of women. Since the effects of aging on muscle fatigability are most evident at higher speeds of movement [], we might also have observed an effect of dietary NO_3_^-^ on fatigability had we tested at a higher velocity of knee extension. Similarly, it is possible that NO_3_^-^ supplementation might enhance recovery from fatigue induced by other types and/or durations/intensities of exercise.

## CONCLUSION

In summary, we have assessed the effects of acute NO_3_^-^ intake on maximal isokinetic knee extensor torque during and after high intensity dynamic exercise in older women. Despite resulting in significant elevations in plasma NO_3_^-^ and NO_2_^-^ levels, and hence presumably in NO levels, NO_3_^-^ supplementation had no effect on muscle fatigability or recoverability under these conditions.

## Data Availability

All data produced in the present study are available upon reasonable request to the author

## FUNDING

This work was supported by the Office of the Vice Provost for Research at Indiana University Purdue University Indianapolis and by the National Institutes of Health (grant numbers R21 AG053606 to ARC, K23 DK102824 to RNM, P30 AR072581 to Sharon Moe, and UL1 TR002529 to Anantha Shekhar). The contents of this article are solely the responsibility of the authors and do not necessarily represent the official view of the National Institutes of Health.

## DISCLOSURE STATEMENT

The authors have nothing to disclose.

## Notes

### Competing Interest Statement

The authors have declared no competing interest.

### Clinical Trial

NCT03595774

### Author Declarations

The Institutional Review Board of the Human Subjects Office of Indiana University gave eithical approval for this work

